# Harnessing Artificial Intelligence for Accurate Diagnosis and Radiomics Analysis of Combined Pulmonary Fibrosis and Emphysema: Insights from a Multicenter Cohort Study

**DOI:** 10.1101/2025.01.20.25320811

**Authors:** Shichen Zhang, Hongqiu Wang, Haiyun Tang, Xiaoqin Li, Nian-Wei Wu, Qin Lang, Bo Li, Hongbin Zhu, Xin Chen, Kaishi Chen, Baosong Xie, Aiyuan Zhou, Chunheng Mo

**Affiliations:** The Key Laboratory of Birth Defects and Related Diseases of Women and Children of MOE, State Key Laboratory of Biotherapy, West China Second University Hospital, Sichuan University, Chengdu 610041, China; School of Engineering, Westlake University, Hangzhou 310030, China; Department of Systems Hub, The Hong Kong University of Science and Technology (Guangzhou), Guangzhou, China; Department of Radiology, Xiangya Hospital, Central South University, Changsha, Hunan, China; Department of Respiratory Medicine and Critical Care Medicine, Fujian Provincial Hospital, Fuzhou University Affiliated Provincial Hospital, Fuzhou 350001, China; Department of Pulmonary and Critical Care Medicine, West China Hospital, Sichuan University, Chengdu 610041, China; Department of Pulmonary and Critical Care Medicine, Sichuan Provincial People’s Hospital, University of Electronic Science and Technology of China, Chengdu 610000, China; Department of Physiology, West China School of Basic Medical Sciences and Forensic Medicine, Sichuan University, Chengdu 610041, China; Department of Respiratory and Critical Care Medicine, Xiangya Hospital, Central South University, Changsha, Hunan 410011, China

**Keywords:** CPFE, deep learning, radiomics, multicenter research

## Abstract

Combined Pulmonary Fibrosis and Emphysema (CPFE), formally recognized as a distinct pulmonary syndrome in 2022, is characterized by unique clinical features and pathogenesis that may lead to respiratory failure and death. However, the diagnosis of CPFE presents significant challenges that hinder effective treatment. Here, we assembled three-dimensional (3D) reconstruction data of the chest High-Resolution Computed Tomography (HRCT) of patients from multiple hospitals across different provinces in China, including Xiangya Hospital, West China Hospital, and Fujian Provincial Hospital. Using this dataset, we developed CPFENet, a deep learning-based diagnostic model for CPFE. It accurately differentiates CPFE from COPD, with performance comparable to that of professional radiologists. Additionally, we developed a CPFE score based on radiomic analysis of 3D CT images to quantify disease characteristics. Notably, female patients demonstrated significantly higher CPFE scores than males, suggesting potential sex-specific differences in CPFE. Overall, our study establishes the first diagnostic framework for CPFE, providing a diagnostic model and clinical indicators that enable accurate classification and characterization of the syndrome.

## Introduction

Combined Pulmonary Fibrosis and Emphysema (CPFE) is a distinct clinical syndrome characterized by the simultaneous presence of pulmonary fibrosis and emphysema, primarily identified through computed tomography (CT). CPFE patients typically exhibit severe dyspnea, hypoxemia, and a high prevalence of comorbidities, including pulmonary hypertension, contributing to significant morbidity and mortality. Despite its clinical importance, CPFE was only recently recognized as a distinct disease entity in 2022^1–7^, reflecting its complex nature and the difficulty of delineating it from other lung conditions, such as chronic obstructive pulmonary disease (COPD) and idiopathic pulmonary fibrosis (IPF)^8–18^. This delayed recognition underscores the challenges in understanding CPFEs pathogenesis, clinical course, and effective diagnostic strategies.^2,19^

The diagnosis of CPFE relies heavily on imaging. Emphysema is defined as well-demarcated areas of low attenuation, delimited by a very thin wall (≤1 mm) or no wall, and involving at least 5% of the total lung volume, lung fibrosis can be of any subtype^20,21^. However, distinguishing CPFE from other diseases with overlapping imaging and clinical features remains a major challenge, even for experienced radiologists. Current approaches lack standardization, and the variability in radiological interpretation often contributes to misdiagnoses. Furthermore, most existing studies on CPFE are limited to small cohorts or single-center datasets^22–25^. The absence of a systematic framework for classifying and quantifying CPFE further hampers efforts to improve patient outcomes.^26–28^

Advances in artificial intelligence (AI), particularly deep learning (DL), offer unprecedented opportunities to enhance diagnostic accuracy and standardization in medical imaging^29–31^. DL models have demonstrated remarkable success in identifying complex patterns within radiological data, facilitating the diagnosis of lung diseases such as COPD, IPF, and lung cancer. However, no AI-driven approach has been specifically developed for CPFE, a condition that uniquely combines features of both emphysema and fibrosis.

Here, we present CPFENet, the first deep learning model specifically designed to classify CPFE, COPD, and control cases (normal lungs) based on 3D chest CT data. This model was developed and validated using a large, multicenter dataset (2836 cases) encompassing patient samples from Xiangya Hospital, West China Hospital, and Fujian Provincial Hospital, making it one of the most comprehensive studies on CPFE to date. CPFENet achieves superior diagnostic performance (83.3% for accuracy) compared to both junior and senior radiologists (around 71% accuracy for senior doctors and around 53% accuracy for junior doctors), demonstrating its potential to address the challenges of CPFE diagnosis in clinical practice. Beyond classification, we further explore the interpretability of CPFENet by integrating its extracted features with radiomics data. This integration enables the development of a novel clinical metric, the CPFE score, which provides a quantitative assessment of CPFE severity.

In summary, this study represents the first multicenter, systematic effort to develop an AI-based diagnostic framework for CPFE. By combining deep learning and radiomics, our work provides a robust tool for the classification and characterization of CPFE, addressing critical gaps in its diagnosis and evaluation. The methods and findings presented here establish a foundation for future research in CPFE and set a benchmark for AI-assisted diagnostic tools in complex lung diseases.

## Results

### 1.1 Patient Characteristics

Between April 2021 and March 2024, we conducted a multicenter study, collecting imaging data from Xiangya Hospital of Central South University, Fujian Provincial Hospital, and West China Hospital of Sichuan University. Following the exclusion of patients with other lung diseases, such as lung cancer, comorbid thoracic conditions, a history of thoracic surgery, and those with significant image artifacts, a total of 2836 patients were included in the study cohort.

We engaged a panel of expert radiologists and pulmonologists to meticulously review the CT images and other relevant clinical data of these patients, thereby rendering a comprehensive diagnosis and assigning diagnostic labels (as depicted in Figure 1a). Subsequent to the acquisition of the 3D CT dataset for CPFE from the three distinct medical centers, a consensus panel of physicians was convened to establish a standardized diagnostic protocol.

**Figure 1:**
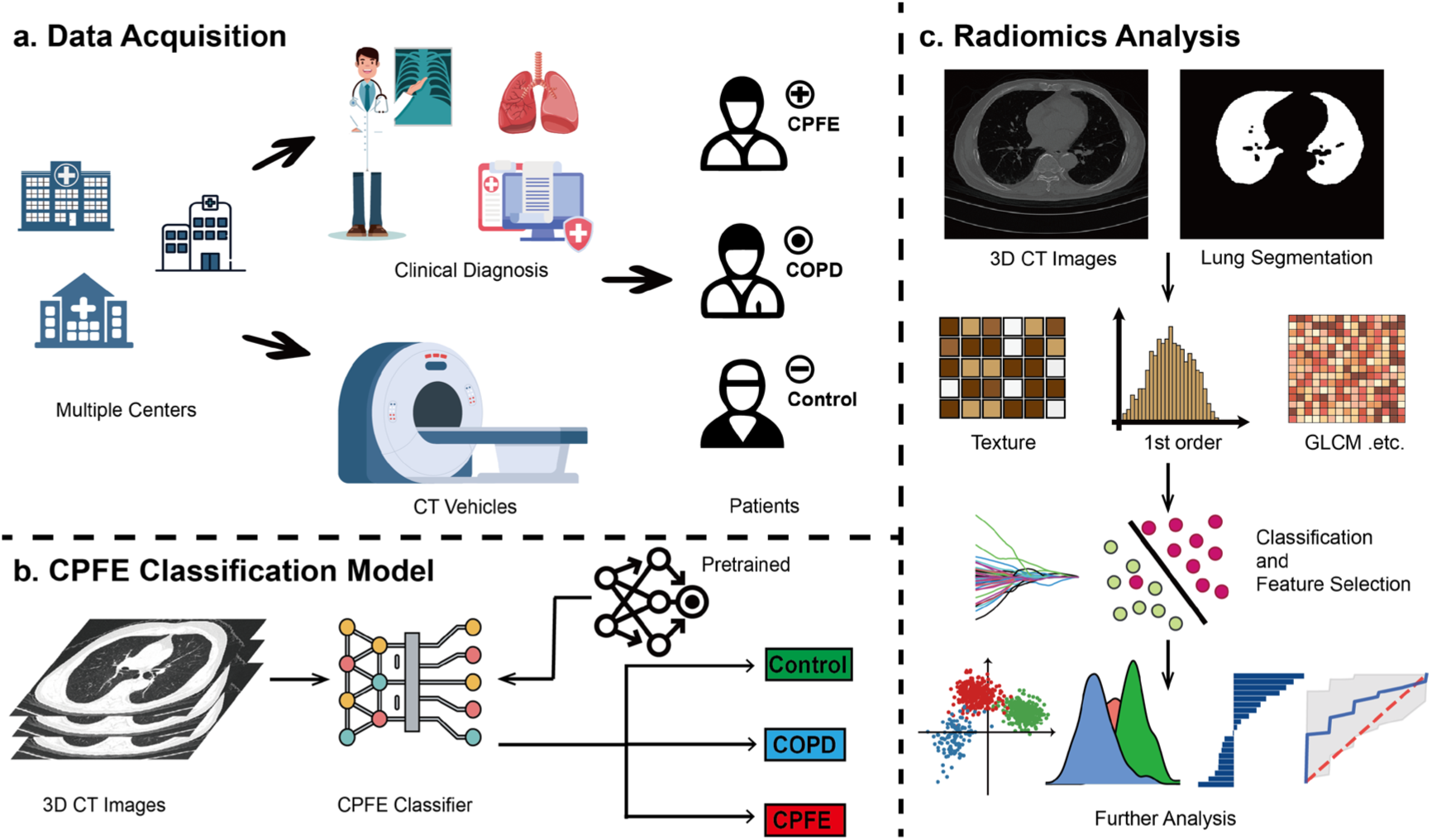
Workflow of the whole study, firstly, in plot a, we acquire high resolution CT (HRCT) data from three clinical centers and we welcome three independent professional radiologists to diagnose the diseases and divided them into three labels including control (normal lungs), COPD and CPFE. Next, we design a deep learning model, which is shown in plot b to use artificial intelligence to affiliate the diagnosis. Finally, in plot c, radiomics are employed to construct the index to represent the disease to find relationship between disease and other clinical influencing factors.

Given the relative low prevalence of CPFE and the lack of a singular diagnostic criterion, we harmonized the diagnostic criteria for both COPD and CPFE. Consequently, a total of 405 CT cases corresponding to CPFE, 1175 cases of COPD, and 1256 control cases (normal lungs) were meticulously curated for the subsequent analysis (as illustrated in Figure 2). This rigorous data curation process ensured the integrity and relevance of the datasets, thereby facilitating a robust investigation into the diagnostic nuances of CPFE and COPD.

**Figure 2:**
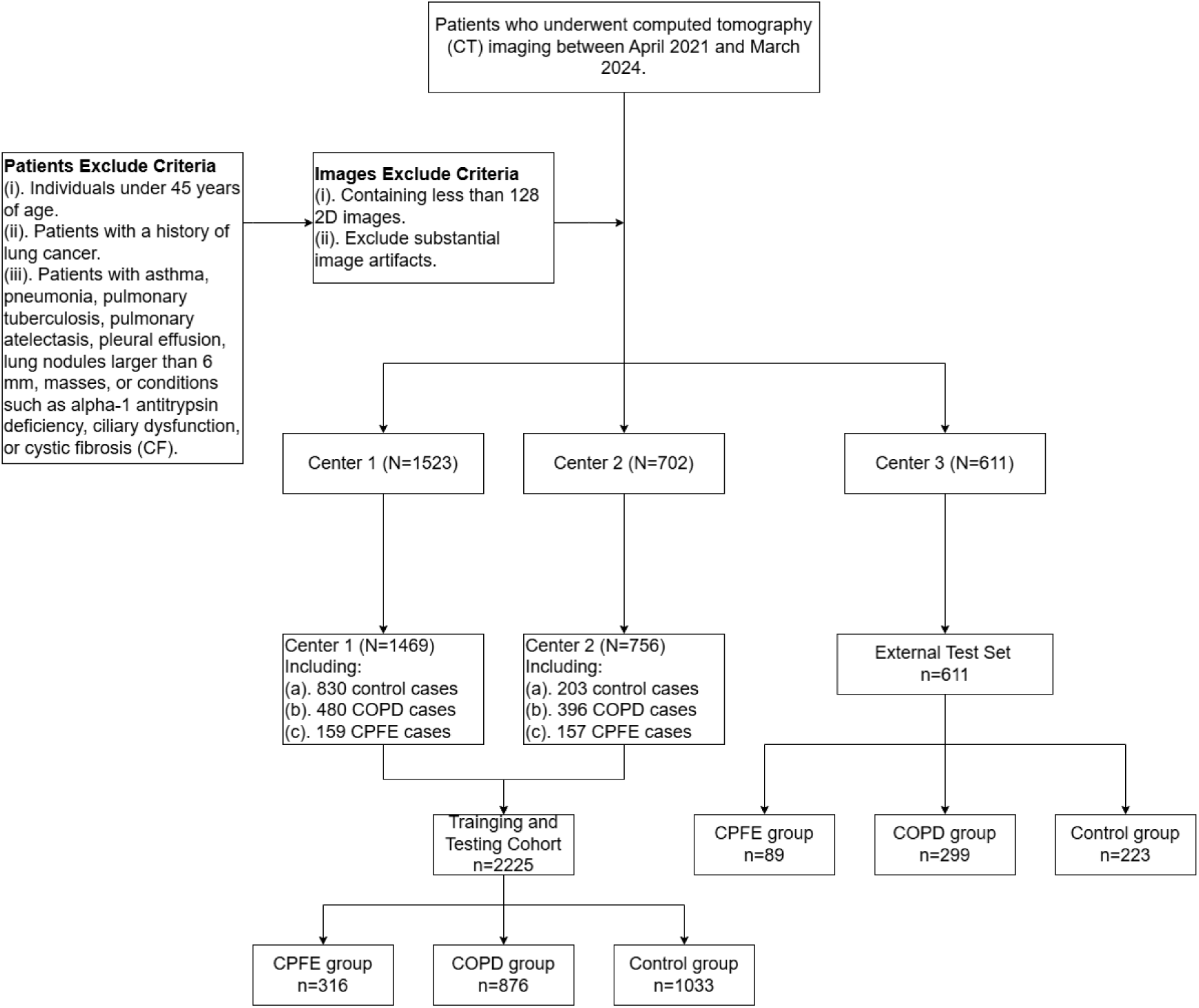
Patients selection flowchart. COPD, Chronic Obstructive Pulmonary disease; CPFE: Combined Pulmonary Fibrosis and Emphysema; Control, normal lung.

### 1.2 Auxiliary diagnosis deep learning model-CPFENet

Despite the involvement of experts in assigning accurate labels to the CT data, inherent variabilities in treatment protocols and disease pathogenesis, along with the predominant reliance on physicians subjective interpretation of CT images for hospital -based diagnoses, have given rise to inconsistencies in disease identification. This issue is particularly pronounced in less technologically advanced medical facilities and among less experienced physicians. Furthermore, the paucity of large-scale datasets poses a significant impediment to conducting comprehensive studies on CPFE. Against this backdrop, we endeavored to develop a deep learning model to aid clinicians in the diagnosis of CPFE and to distinguish it from COPD.

Specifically, we devised a deep learning model that harnesses 3D CT data, with a focus on discerning both global and local lesion characteristics pertinent to COPD and CPFE (as illustrated in Figure 1b). The model, termed CPFENet, capitalizes on the feature extraction prowess of Convolutional Neural Networks (CNNs)^32^ and incorporates a self-attention mechanism to meticulously scrutinize each pixel region. In its architecture, we utilized a multi-layer Swin Transformer^33^ block as the transformer component and a residual cascade to constitute the CNN module. These modules process 3D CT data and amalgamate their extracted features via a sophisticated fusion layer. For the training regimen, we adopted a transfer learning approach, wherein individual model modules were pre-trained prior to fine-tuning their parameters within the context of the fully assembled model (as delineated in Figure 3). This methodological framework was designed to optimize the models performance and generalizability, thereby enhancing its diagnostic efficacy in the clinical setting.

**Figure 3:**
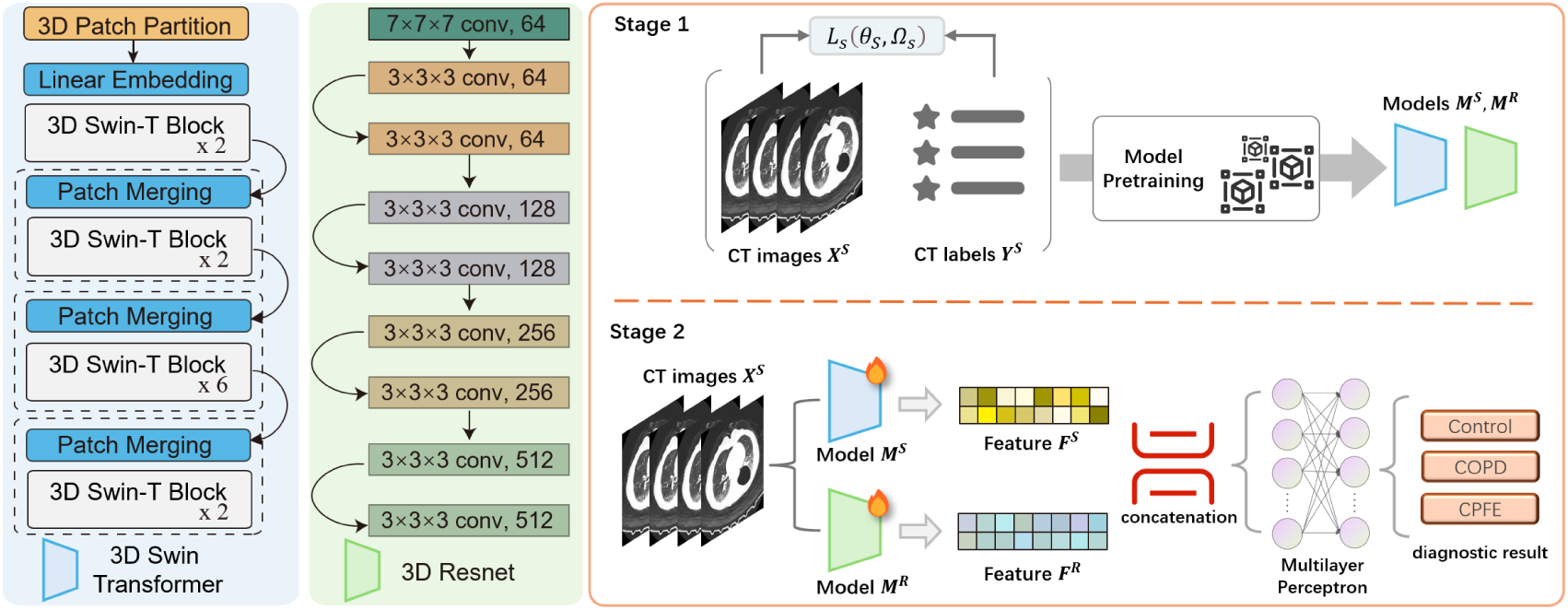
the model overview of the CPFENet. The model employed transferring learning to catch the local and global information by utilizing transformer and CNN module in the 3D CT images and fuse the features to diagnosis.

Following the training of the CPFENet model utilizing the designated training dataset, we sought to evaluate its performance in classifying COPD and CPFE relative to other established deep-learning models. To this end, we constructed a series of benchmark models, encompassing various configurations of 3D ResNet to represent CNN-based architectures and the Swin Transformer to exemplify transformer-based models. Upon analyzing the results derived from both internal and external validation datasets, our CPFENet model demonstrated superior performance across multiple evaluation metrics.

Specifically, for the test dataset, the Swin Transformer model attained an accuracy of approximately 80%, while the ResNet models achieved accuracies ranging from 85% to 87%. In contrast, our CPFENet model achieved an accuracy of 90%. In the external validation dataset, CPFENets performance was further underscored by its AUC and PR values, which surpassed those of the comparator models, reaching 0.94 and 0.9, respectively (as depicted in Figures 4a and 4b, supplementary figure 1a). The accuracy of CPFENet was 0.82, outperforming the 0.78 achieved by ResNet and the 0.71 by Swin-Transformer (as shown in Figure 4c). Collectively, these findings indicate that our work substantially enhances the overall classification accuracy and diagnostic precision for CPFE when compared to other baseline models.

**Figure 4:**
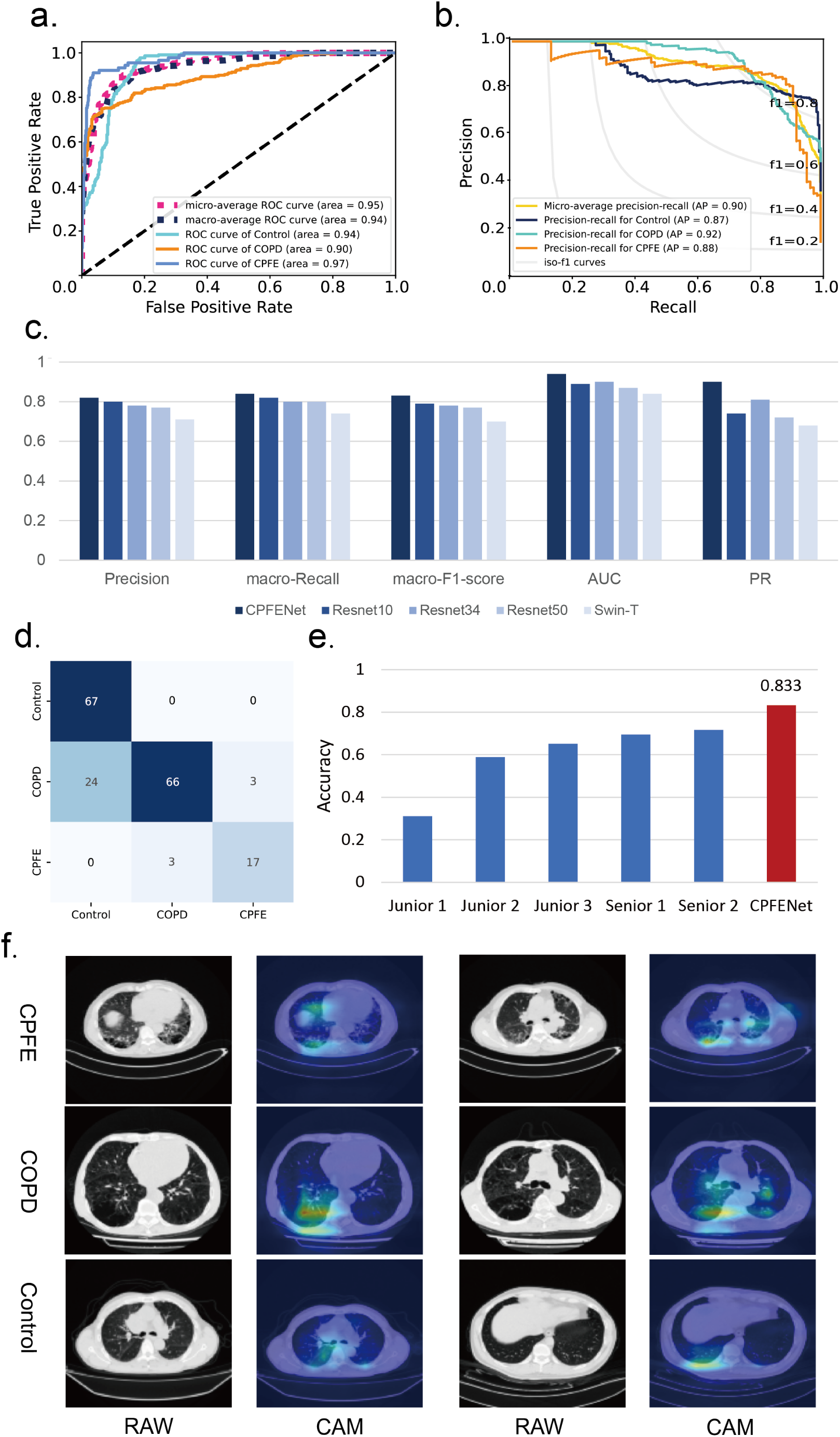
the performance result of the CPFE model. To be specific, the subfigure a and b shows the ROC curve and PR curve of the CPFENet. The subfigure c shows the performance comparing to other baseline models, concrete evaluation indicators include precision, macro-recall, macro-F1 score, AUC and PR results. The subfigure d shows the confusion matrix of the CPFENet prediction result in the diagnostic evaluation dataset. The figure e shows the accuracy of the senior and junior doctor in the diagnostic evaluation dataset. The subfigure f shows the visualization CAM plots of the model intermediate layer parameters accompanying with the raw CT images.

To ascertain the clinical viability of our model, we engaged a panel of experts from West China Hospital and Xiangya Hospital to conduct comparative analyses. A random selection of 180 CT images from the external validation cohort was subjected to diagnostic evaluation. Physicians were stratified into two groups: senior (n=2) and junior (n=3) doctors, based on their hierarchical positions and cumulative experience within the imaging department. Our analyses revealed a significant correlation between the success rates of CPFE and COPD diagnoses and the level of expertise of the radiologists. Junior radiologists demonstrated an accuracy rate of approximately 53%, whereas senior radiologists achieved an accuracy rate of 71%. In stark contrast, CPFENet exhibited an accuracy rate of 83.3% (as shown in the figure 4d and figure 4e), thereby surpassing the diagnostic proficiency of physicians across all experience levels and affirming the models robust stability and precision. The detailed diagnostic outcomes for each physician are presented in Supplementary Table 1.

To ascertain the models ability to discern salient features within the 3D CT images, we implemented a class activation mapping (CAM) process (as illustrated in Figure 4f). The resultant CAM visualizations revealed that our model accurately localized the lesion areas associated with COPD and CPFE, thereby validating its capacity to identify critical diagnostic features within the imaging data. This capability is instrumental in facilitating accurate and timely diagnoses, which is of paramount importance in the clinical management of these pulmonary conditions.

In summary, CPFENet demonstrates a marked capacity to accurately discern CPFE from 3D CT data, representing a substantial enhancement over traditional physician-based diagnoses in clinical contexts. We posit that CPFENet holds considerable promise for augmenting practical clinical applications and may serve as a transformative catalyst in the evolution of CPFE diagnostics, potentially revolutionizing the approach to disease identification and management in this domain.

### 1.3 CPFE score acquisition based on radiomics features

Indeed, in conjunction with leveraging 3D CT scans to facilitate deep learning - assisted diagnosis, our objective extends to conducting an exhaustive analysis of the imaging characteristics of CPFE and their correlation with pertinent clinical indicators. Our deep learning model has instilled confidence in the feasibility of diagnosing CPFE through the analysis of 3D CT images, thereby prompting our conviction that the extraction of image - based features can aid in the construction of a comprehensive scoring system for the disease. Consequently, we aspire to quantify the disease progression by employing radiomic features, with the ultimate aim of defining a CPFE - specific clinical metric, herein designated as the CPFE score.

However, acknowledging the interpretability challenges inherent to deep learning models and the non - uniform distribution of features within such data, we have elected to utilize radiomic features for a more robust and interpretable index evaluation. To this end, we harnessed data from the Xiangya Hospital Center, integrating clinical indicators and patient - specific information to segment lung images and extract radiomic features. From the 780 3D CT images analyzed, we amassed over 1300 radiomic feature data points.

Due to the noise features and other problems in radiomics, the use of unsupervised clustering methods including PCA, TSNE and UMAP (as shown in supplementary figure 2,3,4) will produce poor results. We utilized Linear Discriminant Analysis (LDA)^34^ for visualization, we observed that COPD cases were positioned as an intermediary group between the control and CPFE cohorts within the latent space (as depicted in Figure 5a). This observation aligns with the established medical understanding of disease progression and underscores the capacity of radiomic features to effectively represent the disease spectrum. Such findings not only validate the utility of radiomic features in disease characterization but also highlight their potential as a valuable adjunct in the clinical assessment of CPFE.

**Figure 5:**
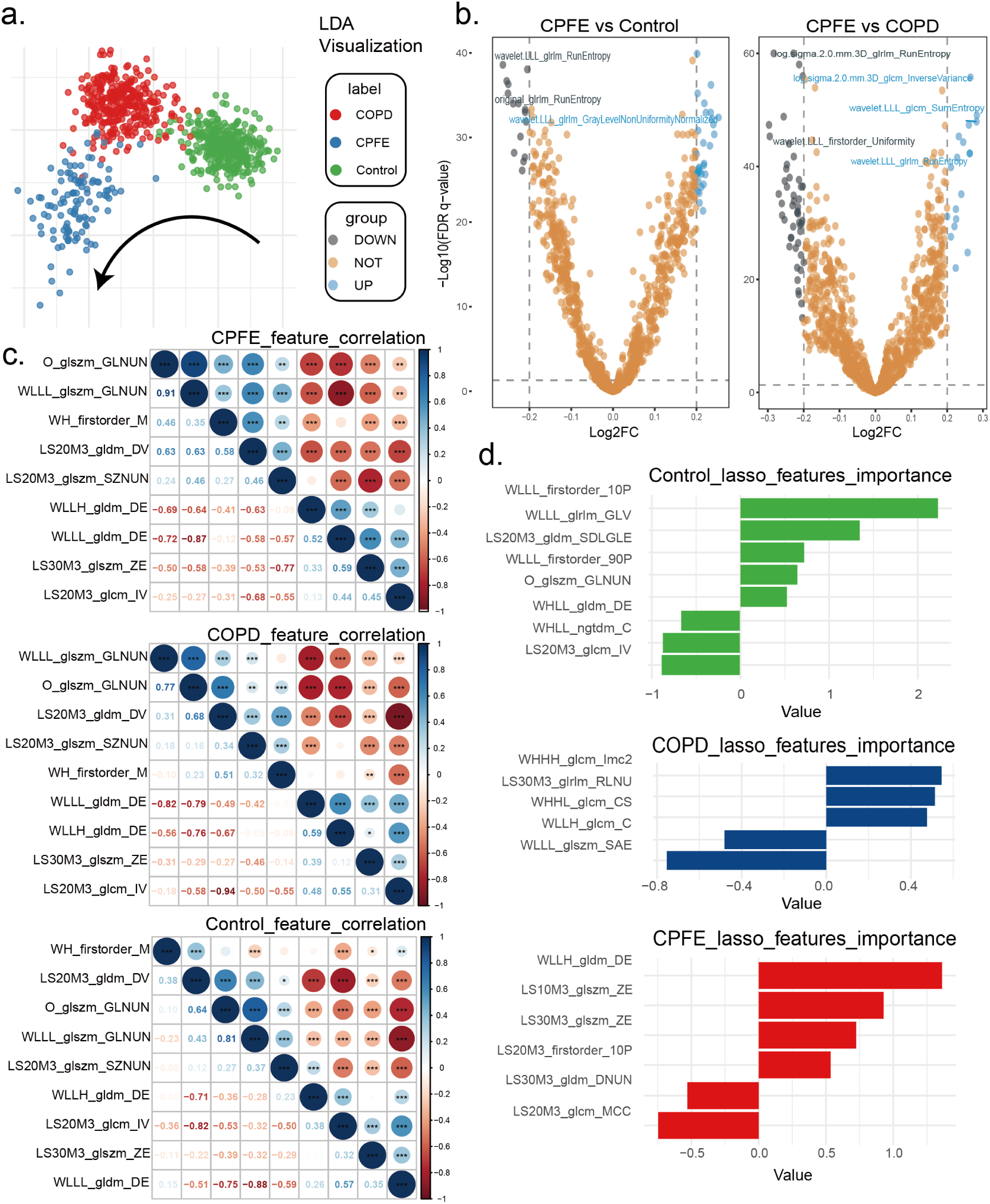
the radiomics analyze result and important radiomics feature selection. Plot a shows the LDA visualization of all radiomics features. Plot b shows the volcano plot of the differentially expression features in CPFE vs Control group (normal lungs) and CPFE vs COPD group. Plot c represents the correlation between the selected radiomics features in the CPFE, COPD and Control group. Plot d shows the feature importances calculated by the lasso model in the Control, COPD and CPFE groups.

Subsequently, the identification of salient radiomic features became imperative for advancing our research. Following the initial step of screening for differentially expressed features (as illustrated in Figure 5b), we employed Lasso regression for feature selection (as depicted in Figure 5d). This rigorous analytical process led to the identification of nine radiomic features that were deemed critical for the construction of the CPFE score. A correlation analysis of these features across the three groups revealed distinct correlation patterns, which provided a robust foundation for the definition of the CPFE score (as shown in Figure 5c).

Consequent to the radiomic feature analysis, we selected the Support Vector Machine (SVM)^35^ model, which exhibited the highest covariance, as the basis for the CPFE score. This model demonstrated an impressive Area Under the Curve (AUC) of 97% (as presented in Figure 6a), a metric that underscores the high degree of complementarity among the radiomic features and their collective ability to capture the imaging - based commonalities characteristic of CPFE. The contribution graph of the various features to the score further substantiated the necessity and efficacy of employing diverse feature extraction methodologies (as shown in Figure 6b).

**Figure 6:**
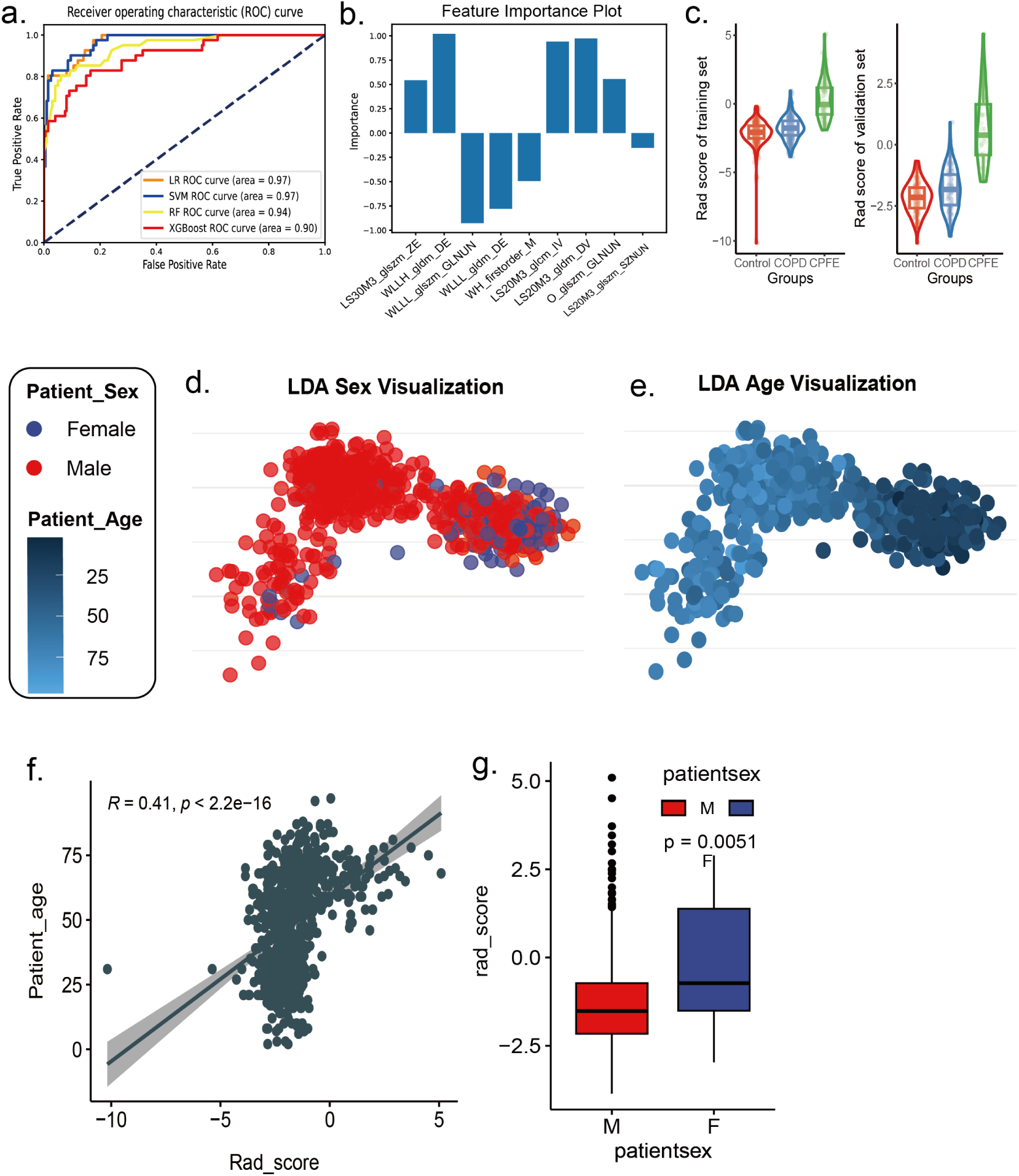
the calculation of the CPFE score and clinical index correlation calculate. Plot a shows the ROC curve of different machine learning models in the selection of the CPFE score. Plot b shows different importance of the radiomics in the calculation of CPFE score. Plot c is the violin plot of the distribution of the calculated CPFE score in the training set (left) and validation set (right). Plot d is the LDA dimensionality reduction plot with the sex label. Plot e is the LDA dimensionality reduction plot with the age label. Plot f is the correlation plot between the age and the CPFE score. Plot g is the boxplot of the CPFE score in different sex group. Plot h is the boxplot of the CPFE score in different sex and age group, which shows the significant difference in the groups of patients under age 70.

Upon examining the distribution of CPFE scores within both the training and test datasets, a consistent pattern emerged wherein the COPD group exhibited higher scores than the control group (as illustrated in Figure 6c). This observation is in concordance with the prevailing hypothesis that CPFE may evolve directly from COPD, thereby lending scientific credence to the validity of the CPFE score as a diagnostic and prognostic tool.

To further unravel the underlying mechanisms of the disease and to enhance our understanding of its progression, we embarked on a comprehensive joint analysis incorporating the CPFE score with other pertinent clinical and radiological indicators. This integrative approach is anticipated to provide deeper insights into the pathophysiology of CPFE, potentially paving the way for more targeted and effective therapeutic interventions.

### 1.4 Clinical indicators related to CPFE disease

The CPFE score, which is derived from radiomic features, has exhibited robust reliability and scientific validity, as evidenced by its high predictive accuracy and consistent distribution across diverse patient groups. In an effort to discern the underlying patterns of CPFE disease, we performed an analysis to explore if the age and gender of the patients are associated with the CPFE score. Utilizing dimensionality reduction plots (as depicted in Figures 6d and 6e), we observed that different patient cohorts may exhibit discernible patterns in these demographic indicators. Consequently, we are intrigued by the prospect of correlating the CPFE score with clinically relevant parameters to glean novel insights into CPFE from an indicator - based perspective.

Our preliminary analysis centered on the correlation between the CPFE score and patient age, which revealed a moderate correlation with a Pearson correlation coefficient of 0.41 (as shown in Figure 6f). Given the higher prevalence of COPD in middle-aged and elderly individuals, statistical analyses, including dimensionality reduction and T-tests, revealed no significant differences between the COPD and CPFE groups (*P* > 0.05). Thus, the correlation with age primarily corroborates the demographic characteristics of COPD in older populations. Moreover, a gender - specific analysis indicated that CPFE scores were significantly elevated in females diagnosed with CPFE, suggesting potential sex-specific differences in CPFE (as shown in Figure 6g and Figure 6h).

## Methods

### 2.1 Data acquisition and preprocessing

This retrospective, multicenter study received approval from the local institutional review boards of all participating centers, with the requirement for informed consent waived. The training and internal test datasets were sourced from patients at Xiangya Hospital of Central South University and Fujian Provincial Hospital, while the external test dataset comprised patients from West China Hospital of Sichuan University.

Following consultations with radiologists, we considered including 2836 patients who underwent lung CT examinations between April 2021 and March 2024 in this study, based on the cohort data from the three centers.

The imaging manifestations of COPD include emphysema and chronic bronchitis. Emphysema is characterized by low - density areas devoid of visible walls and is categorized into three subtypes: centrilobular, paraseptal, and panlobular. Chronic bronchitis is typified by bronchial wall thickening.

The imaging manifestations of CPFE are characterized by the presence of emphysema in the upper lung zones and fibrosis in the lower lung zones^3^. Interstitial fibrosis is defined by areas of increased parenchymal attenuation, presenting as reticular opacities and/or ground – glass opacities, variable degrees of honeycombing, and/or traction - induced bronchiectasis.

Ethical approval for this study was granted by the institutional review boards of Xiangya Hospital of Central South University, Fujian Provincial Hospital, and West China Hospital of Sichuan University, and the study was conducted in strict adherence to the ethical standards set forth in the 1964 Declaration of Helsinki. Given the retrospective design of the study, the need for informed consent was waived.

For the preprocessing of the 3D CT data, the SimpleITK (sitk) Python package was utilized. The ResampleImageFilter method was employed to uniformly resample the image voxels to a size of [1.0, 1.0, 1.0] mm³, and the CT image direction was standardized. The window width was set to 1300 Hounsfield units (HU) and the window level to -450 HU. Subsequently, the image grayscale values were normalized, and the image dimensions were uniformly resized to 128 × 128 × 128 pixels to serve as the standardized input for the model.

### 2.2 CPFENet Model construction

In order to solve the characteristics of CPFE disease such as difficult diagnosis, easy confusion and low frequency, we designed a multi-stage deep learning-assisted diagnosis model CPFENet that combines CNN and Transformer framework using transfer learning knowledge. To address the challenge of distinguishing between COPD and CPFE, we categorized the data into three groups: Control, COPD, and CPFE. This approach aims to identify differences between COPD and CPFE within the models classification of CPFE, thereby enabling accurate segmentation of CPFE. For data allocation, we randomly split the samples into training and testing sets at a ratio of 7:3 to support subsequent model training.

In terms of the specific architecture of the model, since the commonly used deep learning frameworks for medical image processing at this stage are mainly divided into two mainstream models: CNN-based^32^ and Transformer-based^33^, in order to more effectively capture the local effective features and global features of 3D lung image CT, we designed a classification network CPFENet specifically for CPFE diseases, which considers the characteristics of CPFE diseases in the presence of local alveoli and the presence of fibrotic parts in other lung regions. The model was constructed by combining the capture of local information by CNN and the learning ability of Transformer to global features. Specifically, the preprocessed 3D CT data needs to be passed through two sub-modules with ResNet and swin transformer as the main framework to extract the features of different points of interest, and then fuse the learned features and add them to the linear layer for accurate classification. We chose ResNet34^32^ as the framework for the first part feature extraction. For another part, we first performed a 3D patch partition for 3D image segmentation, and then used embedding to encode the position information and put it into four layers of 3D swin-T layers, in which the patch merging module between each layer integrates different position information and inputs it into the swin transformer module, and the number of blocks in different layers is set to 2, 2, 6, 2 respectively. Based on the features of the two modules, the three-classification tasks of CPFE, COPD and Control were carried out by concating and then putting them into the classification layer.

In terms of model training, we first used the parameters pre-trained for ImageNet to train the first stage of the model on the 3D ResNet and 3D Swin-Transformer frameworks to learn different disease features from different perspectives. After training, the model parameters were migrated to the CPFENet model framework for the second stage of tuning to obtain the final accurate classification. In terms of hyperparameter settings, we have a learning rate of 1e-5 for the first and second phases, an epoch set to 200, and a batch size of 32.

### 2.3 Radiological features extraction

In order to perform radiomics studies on CPFE, we first need to perform lung parenchymal segmentation by 3D CT, here we use the SimpleITK^36–38^ tool to segment the lung parenchyma by threshold segmentation combined with seed generation. After excluding the missegmented samples, the relevant lung parenchymal radiomics data were extracted by pyradiomics^39^. We mainly extracted the original features, first-order statistical features and second-order statistical features of 3D CT data, and the second-order statistical features were mainly extracted through log and wavelet filters. In the end, we extracted 1302 radiomics features for subsequent analysis.

### 2.4 CPFE related radiological characteristics selection

In order to be able to study the imaging features associated with CPFE, we hope to be able to extract CPFE-related radiomics features in two ways. The first method is based on the lasso algorithm, we select the appropriate lambda value for the cross-validation method of the standardized radiomics data, and use the λ value to perform the lasso regression for the classification task between the control, COPD and CPFE of the sample, and select the part of the regression shrinkage coefficient greater than 0 as the set screened out by the first method, which helps us to select the features with anti-noise ability and easy to calculate. and facilitate the calculation of subsequent CPFE scores. In the second method, the differential expression characteristics of Control vs CPFE and COPD vs CPFE were screened, and the threshold of log foldchange was 0.2 and the *P* value was less than 0.05. In the end, nine relevant traits were screened and applied to the subsequent calculation of the CPFE score.

### 2.5 CPFE score calculation

In the computation of CPFE scores, we utilized nine selected indicators. To specifically target CPFE, we employed machine learning algorithms, including linear regression, support vector machine (SVM)^35^, random forest^40^, and XGBoost^41^, for classification. We selected the model with the highest accuracy for the determination of the CPFE score. Initially, we randomly assigned data to training and test sets in a 7:3 ratio and performed a grid search for hyperparameter optimization in each model. During training, we implemented five-fold cross-validation. Ultimately, we selected the SVM as the machine learning model and adopted its output as the CPFE score metric.

### 2.6 Clinical value assessment

Upon deriving the CPFE scores, we collected the corresponding age and gender data for each score and performed a T-test on these discrete variables to assess significant differences in CPFE scores across patient subgroups. For continuous variables, we computed the Pearson correlation coefficient to identify clinical indicators correlated with the severity of CPFE, thereby informing clinical diagnosis of the disease.

## Discussion

CPFE poses a significant challenge in clinical diagnostics, distinguished by its chronicity, poor prognosis, and frequent co - morbidities, such as pulmonary hypertension and lung cancer^42–46^. Since the initial description of CPFE in 1948, the medical community has accrued a more profound understanding of this intricate syndrome^3,26^. This condition is characterized by marginal reductions in lung volume and profound impairments in pulmonary diffusion capacity, with pathological features encompassing emphysematous changes, inflammatory responses, and fibrotic processes^47–49^. Despite these advances, the diagnosis of CPFE remains reliant on the expertise of radiologists, highlighting the urgent need for the development of innovative diagnostic adjuncts. In this study, we utilized deep learning and radiomics to conduct a comprehensive analysis of CPFE from 3D CT data, with the objective of enhancing diagnostic precision and efficacy.

Within the scope of this investigation, we amalgamated CPFE 3D CT datasets from three disparate centers, thereby establishing a standardized diagnostic protocol and constructing a deep learning model, CPFENet. CPFENet integrates the feature extraction capabilities and self attention mechanisms of Convolutional Neural Networks (CNNs), in conjunction with multi layer Swin Transformer blocks and residual cascades, to markedly enhance CPFE diagnostic accuracy through transfer learning and subsequent fine - tuning^50,51^. Our model outperformed established benchmarks, including different types of 3D ResNet and Swin Transformer, achieving a 90% accuracy rate and surpassing the diagnostic outcomes of physicians across all levels. In the external validation set, our model attained an accuracy rate of 83%, compared to the accuracy rate of approximately 53% for junior doctors and 71% for experienced doctors, thereby affirming CPFENets robustness and reliability.

Furthermore, we delineated the CPFE score, a clinical metric specifically tailored to CPFE that delineates disease trajectory. By incorporating radiomics features, we extracted over 1,800 feature data points from 780 3D CT images, culminating in the identification of 9 pivotal features. These 9 higher-order statistical features were calculated by different filters and analyzed to reflect the main differences in CT images of CPFE patients. The SVM model, with its high covariance outcomes, substantiates the synergistic potential of radiomics in capturing CPFEs imaging characteristics. Distributional analysis of CPFE scores revealed elevated COPD scores relative to control groups in both training and testing cohorts, thereby validating the scientific underpinnings of CPFE scores.

Our research also revealed correlations between CPFE and patient age and sex, with women affected by CPFE exhibiting markedly elevated CPFE scores. These insights offer novel perspectives on CPFEs etiology, genetic influences, therapeutic approaches, and preventative strategies. The identification of sex - based disparities is particularly significant, as it may unravel distinct pathological mechanisms of CPFE across genders, which is crucial for the formulation of targeted therapeutics. Although constrained by data limitations and an absence of follow - up data, our study lays the groundwork for preliminary inter - populational disparity analyses in CPFE. Future research should consider conducting sex - specific sequencing of CPFE biopsy samples to elucidate the role of gender in CPFE^52–54^.

In summary, CPFENet and CPFE scores represent novel diagnostic and investigative tools for CPFE, setting the stage for future research endeavors. We are confident that, with technological progress and data accumulation, CPFENet and CPFE scores will assume increasingly pivotal roles in CPFEs diagnosis, treatment, and prevention. These tools will enable a more nuanced comprehension of CPFEs complexities, inform more precise treatment strategies for patients, and ultimately, enhance patient outcomes and prognoses.

## Data Availability

All data produced in the present study are available upon reasonable request to the authors.

## Declarations

The authors declare that they have no competing interests.

## Acknowledgments

This work was supported by the National Natural Science Foundation of China (82200084), the Natural Science Foundation of Sichuan Province (2023NSFSC1456), the Postdoctoral Science Foundation funded project of Sichuan Province (TB2023047), and the Sichuan University postdoctoral interdisciplinary Innovation Fund (0020404153020).

## Author contributions

S.Z., H.W. and C.M. performed the experiments, data analyses, experimental planning, and wrote the paper. H.T., X.L., N.W., Q.L., B.L., H.Z., X.C., K.C., and B.X. performed the experiments. C.M. and A.Z. conceived and designed the work. All authors contributed to the article, read, and approved the final manuscript.

## Supplementary Figures

**Supplementary figure 1:**
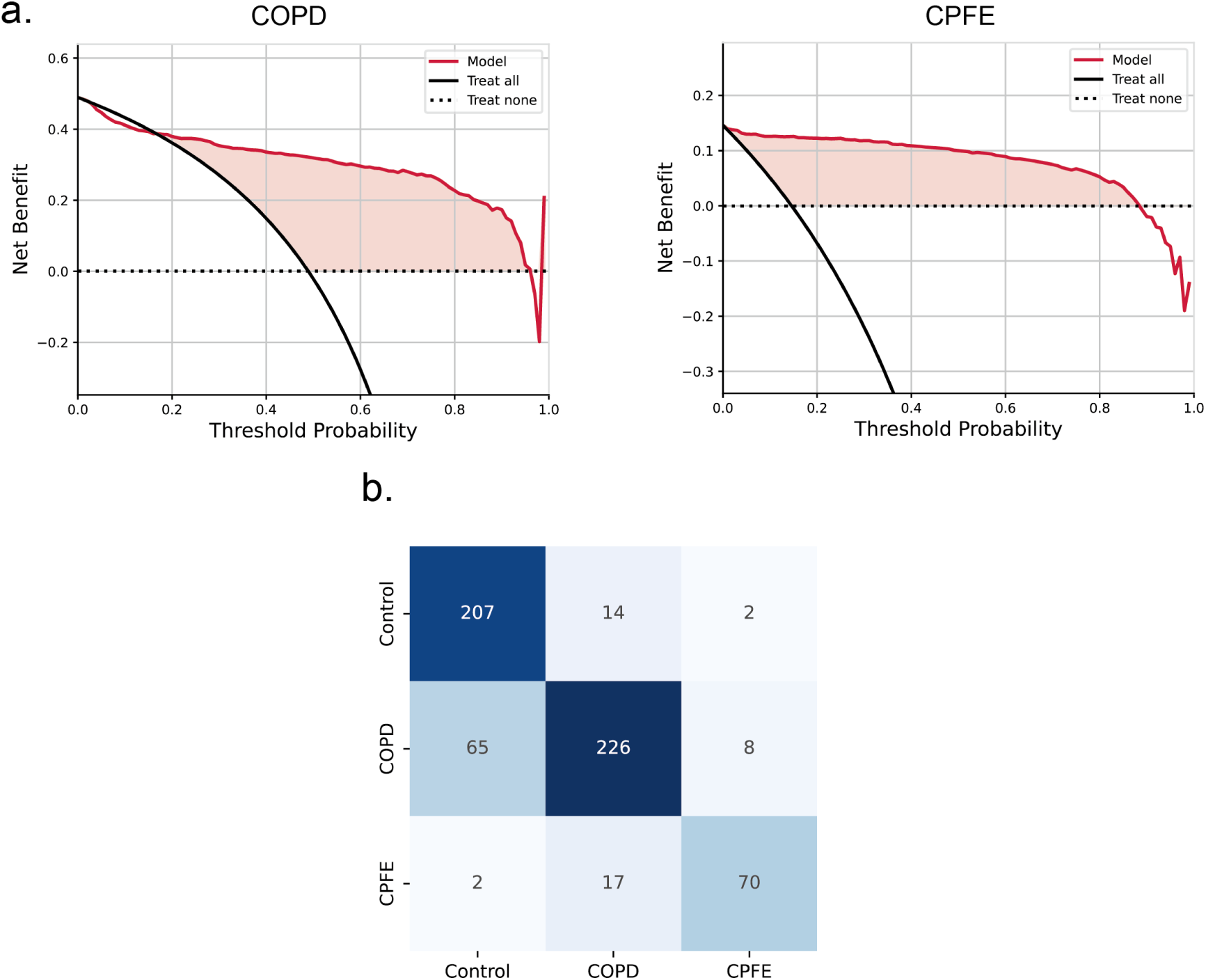
a: the DCA plot of the CPFENet in COPD and CPFE groups, b: the confusion matrix of the CPFENet for the validation dataset.

**Supplementary figure 2:**
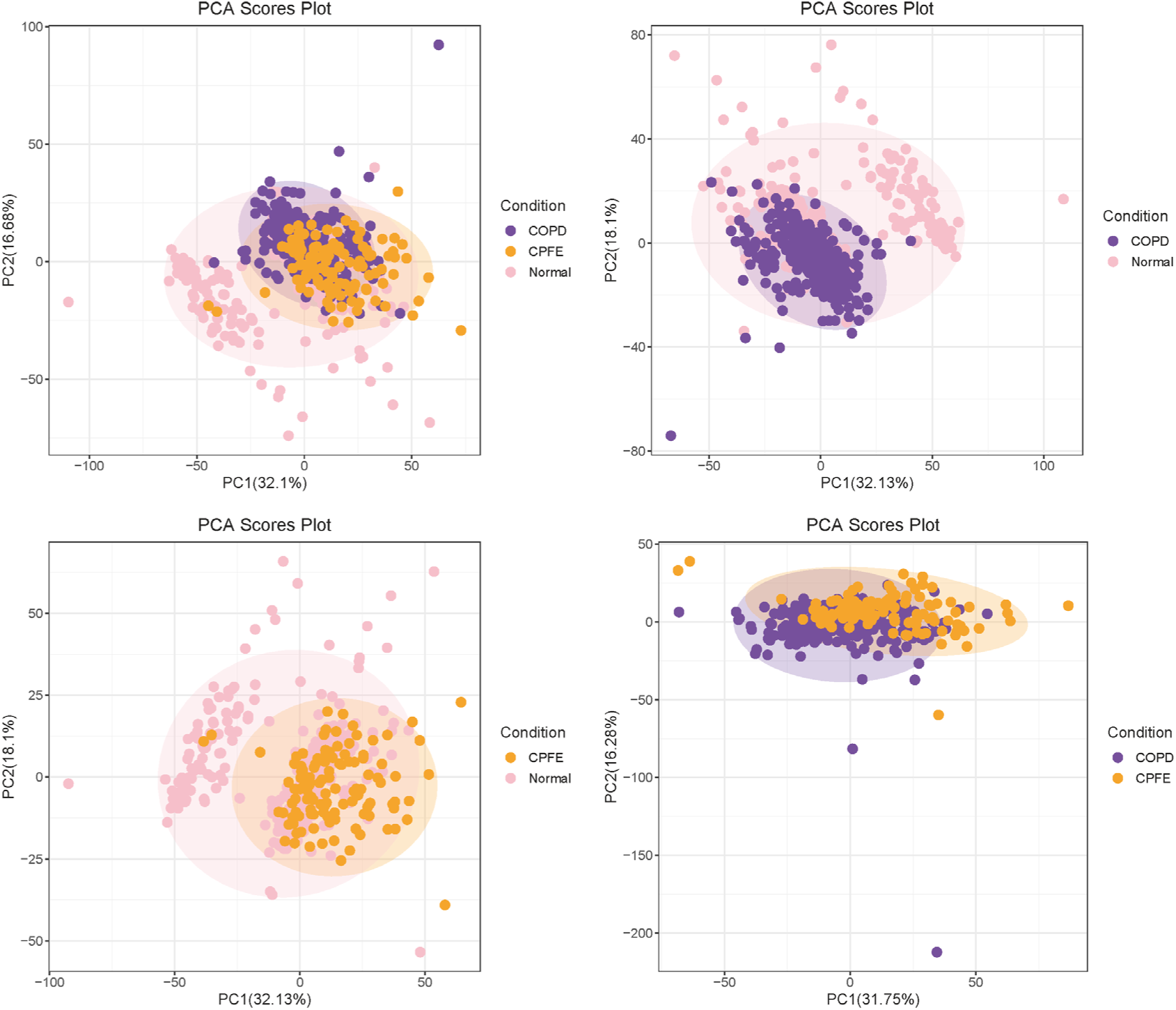
the PCA plot of different groups with radiomics features.

**Supplementary figure 3:**
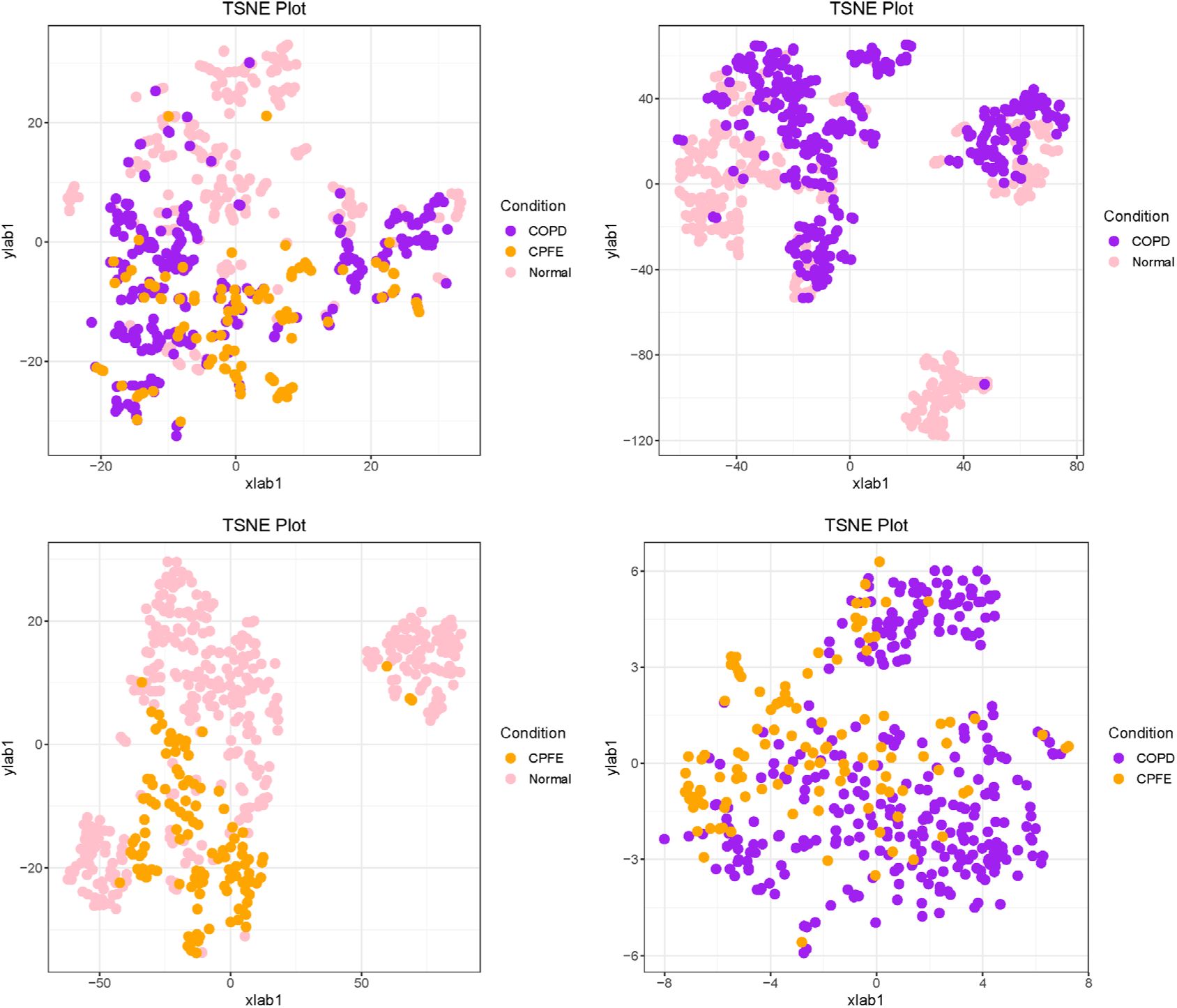
the TSNE plot of different groups with radiomics features.

**Supplementary figure 4:**
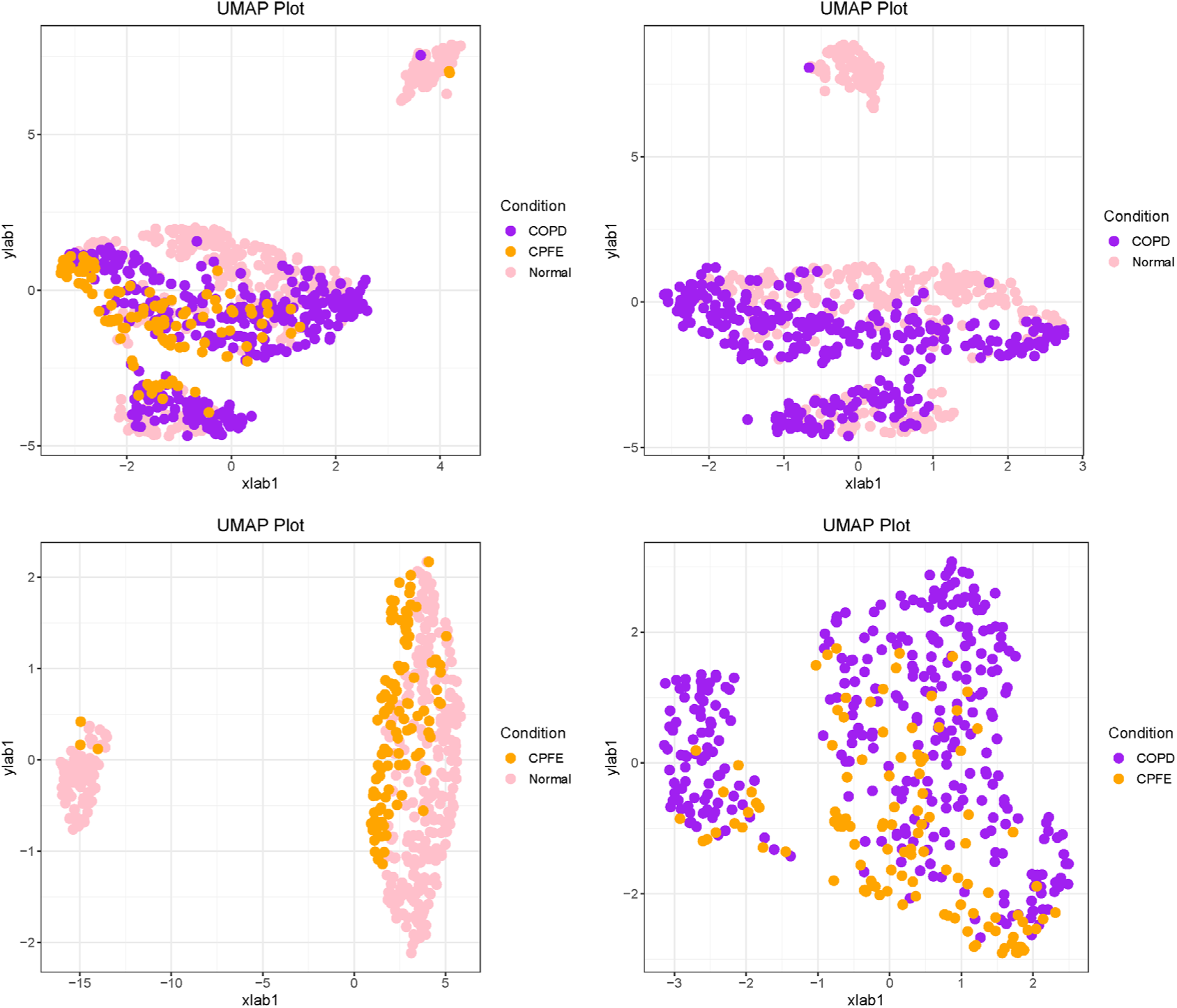
the UMAP plot of different groups with radiomics features.

**Supplementary figure 5:**
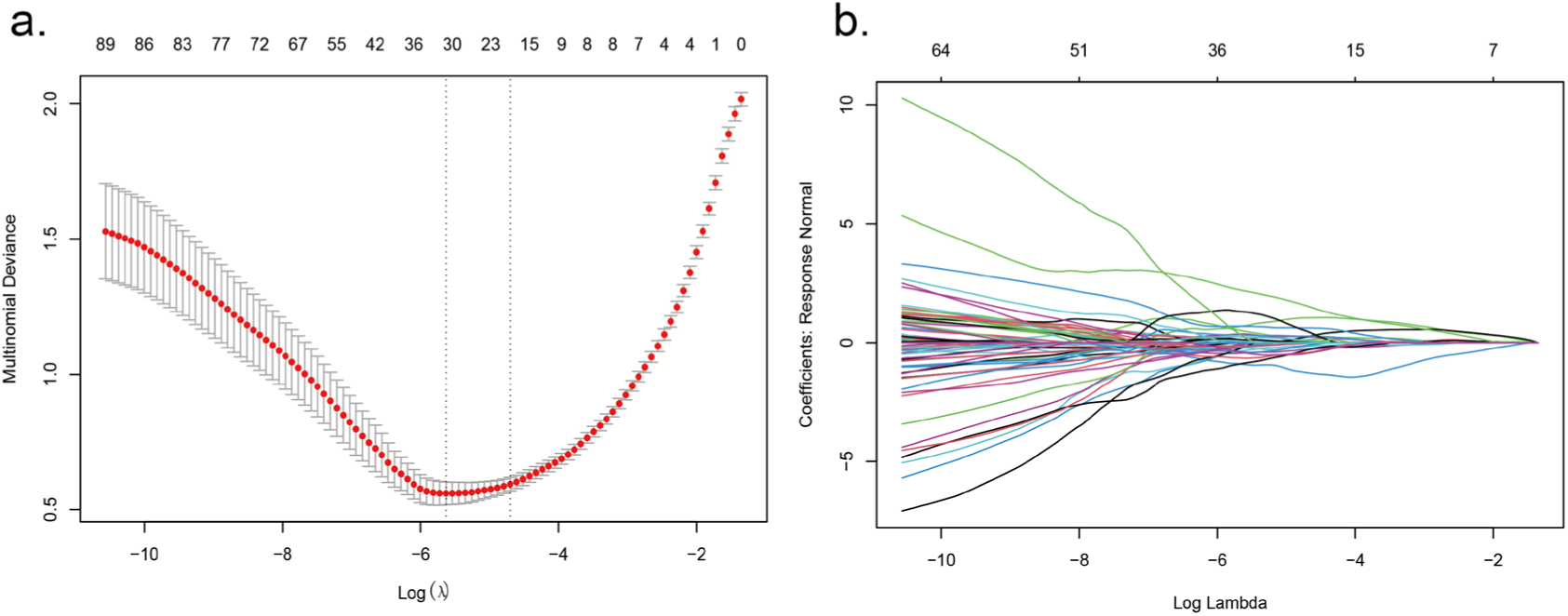
a: the lasso multinomial deviance-log(λ) plot of the lasso model selection, b: the lasso regularization path plot of the selected lasso model.

